# Colorectal Cancer Screening in Adults 45-49: Provider Availability, CT Colonography Access, and Screening Rates

**DOI:** 10.1101/2024.09.25.24314175

**Authors:** Rachel Liu-Galvin, Zhigang Xie, Young-Rock Hong

**Affiliations:** Department of Health Services Research, Management, and Policy, College of Public Health and Health Professions, University of Florida, Gainesville, Florida, USA; Department of Public Health, University of North Florida, Jacksonville, Florida, USA

## Abstract

**Background:** The US Preventive Services Task Force updated colorectal cancer (CRC) screening guidelines in 2021, recommending screening for adults aged 45-49. This study aimed to evaluate CRC screening prevalence among this newly eligible population and examine associations with healthcare provider supply and CT colonography facility availability in 2022.

**Methods:** Using 2022 Behavioral Risk Factor Surveillance System data (n=25,592), we estimated CRC screening prevalence among adults aged 45-49. We examined associations between screening rates and state-level healthcare provider supply using 2021-2022 Area Health Resources File data. Spearman rank-order correlations assessed relationships between provider supply, CT colonography facility availability, and screening prevalence.

**Results:** Overall CRC screening prevalence was 34.5% (95% CI: 33.4%-35.8%). Endoscopic tests were most common (74.9%), followed by stool-based tests (9.3%) and CT colonography (0.5%). Significant variations in screening modalities were observed across sociodemographic factors. Gastroenterology physician supply positively correlated with overall CRC screening prevalence (ρ=0.42, P=.002) and endoscopy screening prevalence (ρ=0.39, P=.005). CT colonography facility availability weakly correlated with CT colonography screening prevalence (ρ=0.18, P=.22).

**Conclusions:** CRC screening rates among newly eligible adults aged 45-49 appear to be suboptimal in 2022. Disparities in screening methods across sociodemographic factors highlight potential access barriers. The association between gastroenterology physician supply and screening rates emphasizes the importance of addressing projected workforce shortages. Targeted efforts are needed to increase CRC screening uptake in this age group and ensure equitable access to screening services.

## Introduction

The US Preventive Services Task Force (USPSTF) recently updated its colorectal cancer (CRC) screening guidelines, to recommend that screening begin at age 45 for average-risk adults (USPSTF, 2021).^1^ This change from the previous recommendation of starting at age 50 aims to address increasing rates of CRC in younger adults and is based on modeling studies showing a favorable balance of benefits and harms.^1–5^ However, comprehensive studies are critical to understanding the impact of this guideline change on screening uptake, particularly among the newly eligible population aged 45-49.

Existing data indicate that CRC screening rates are notably low among adults aged 50-54 (before the guideline update), suggesting that similar challenges may be faced by the 45-49 age group.^6^ This age group has historically been less engaged in preventive health measures, partly due to lower perceived risk and fewer healthcare interactions.^6,7^ Persistent disparities in CRC screening rates are evident across various demographic groups, including race, ethnicity, socioeconomic status, and insurance coverage.^6,8^ These disparities are likely to extend to the newly eligible 45-49 age group, necessitating targeted interventions to ensure equitable access to screening.

Furthermore, the availability and distribution of healthcare providers, particularly primary care physicians and gastroenterologists, play a crucial role in screening rates.^9–11^ Provider recommendations are a strong predictor of screening behavior, and areas with limited provider access may struggle to achieve high screening rates.^12,13^ Similarly, the availability of advanced screening technologies, such as CT colonography, may influence screening modality choice and overall uptake rates.^14,15^ Given these considerations, there is a need for a comprehensive assessment of the current CRC screening landscape for adults aged 45-49.

The aims of this study were to 1) evaluate the prevalence of CRC screening among adults aged 45-49 years in 2022 among US states following the USPSTF guideline update, 2) examine the associations of state-level health care provider supply (primary care physician and gastroenterology physician supply per 10,000 population of adults aged 45-49 years) and prevalence of CRC screening among adults aged 45-49 years in 2022, and 3) examine the association between state-level CT Colonography screening facility availability and prevalence of CT colonography uptake among adults aged 45-49 years in 2022.

## Methods

### Data source

We used the 2022 Behavioral Risk Factor Surveillance System (BRFSS).^16^ The BRFSS is a continuous national telephone survey of non-institutionalized U.S. adults aged 18 years and above that gathers information about individuals’ health-related risk behaviors and events, chronic health conditions, and use of preventive services.^17^ The survey featured a mandatory core section with standardized health-related questions that all participating states and territories were required to use. Additionally, there was an optional module addressing topics like social determinants of health, sexual orientation, and gender identity, which was implemented only in states that chose to include this data.^18^ Information about the BRFSS’ sample design and weighting procedures are reported elsewhere.^19^ Additionally, we utilized state-level data from the 2021-2022 Area Health Resources File (AHRF), which is maintained by the Health Resources and Services Administration.^20^ The AHRF compiles information from over 50 sources to provide comprehensive data on healthcare resources, population characteristics, and health status measures.^20^ For this study, we extracted and aggregated the state-level data on primary care physicians (PCP) and gastroenterology (GI) specialists. PCPs were defined as non-federal doctors of medicine (MD) and doctors of osteopathy (DO) providing direct patient care in general or family practice, general internal medicine, pediatrics, or obstetrics and gynecology. GI specialists were identified as the number of non-federally employed physicians with a medical specialty in gastroenterology. These state-level measures of healthcare provider availability were matched with BRFSS data for analysis of the relationship between healthcare provider density and colorectal cancer screening rates across states. Because the BRFSS and AHRF data are publicly available and deidentified, the Institutional Review Board at the first author’s institute deemed this research exempt from review. Our study adheres to the Strengthening the Reporting of Observational Studies in Epidemiology (STROBE) guidelines.^21^

### Study population

To align with our study objective, we focused on adults aged 45-49 without a history of CRC who responded to the BRFSS 2022 survey (n=28,512). Using a listwise deletion approach, we then excluded respondents with missing CRC screening data, reducing the sample to 26,333 individuals. Additionally, we removed individuals with missing sociodemographic covariate information, resulting in an analytic sample of 25,592 individuals (weighted n=15,362,966).

### Dependent Variables

The primary outcome we assessed was state-level CRC screening prevalence. This was measured according to the percentage of individuals per state who reported having ever had any modality of CRC screening. Modalities of CRC screening assessed in the 2022 Adult BRFSS were as follows: endoscopy (sigmoidoscopy or colonoscopy), stool-based tests (blood stool test, stool DNA test, or Cologuard test), and Computed Tomography (CT) colonography (also known as virtual colonoscopy).

### Independent Variables

In this study, the estimated CRC screening prevalence was focused on census regions (Northeast, Midwest, South, West) and states (including all 50 states and the District of Columbia). We also included the supply of primary care physicians and gastroenterology physicians per 10,000 population aged 45-49 years per state, which was computed using the number of primary care physicians and gastroenterology physicians per state obtained from the state-level AHRF and BRFSS weighted estimates of the population aged 45 to 49 years in each state. We also included the number of CT colonography screening facilities per state, which was computed according to information on the geographic locations of CT colonography screening facilities across the US provided to us by the American College of Radiology (ACR).^22^ Although the ACR website includes a screening locator tool for CT colonography facilities,^23^ to ensure our data was accurate and up to date, we used a list of facilities provided to us directly by the ACR on July 22^nd^, 2024. Other variables incorporated into the analysis include various individual sociodemographic and health-related characteristics.

### Statistical Analysis

Following the complex sampling design of the BRFSS, we conducted weighted analyses using raking weights provided with the dataset.^24^ We computed descriptive statistics to assess differences in the unadjusted CRC screening prevalence across the four regions of the US (Northeast, Midwest, West, and South), and across all fifty US states. We also compared CRC screening prevalence ratios among states based on census region adjusted for sex, race/ethnicity, marital status, education, income, employment, urban/rural location, health insurance coverage, whether an individual had a usual source of care, smoking status, drinking status, body mass index (BMI), general health, number of chronic conditions, and number of disabling limitations. We compared the prevalence of difference types of screening methods undergone according to the following sociodemographic characteristics: region, sex, race/ethnicity, health insurance coverage, urban/rural location, education, and income. We examined five categories of screening modalities as follows: 1) stool-based tests, 2) endoscopic tests (sigmoidoscopy or colonoscopy), 3) CT colonography, 4) a combination of endoscopic tests and stool-based tests or CT colonography, or 5) a combination of stool-based tests and CT colonography. Categories 1), 3) and 5) were considered non-invasive screening methods while categories 2) and 4) were considered invasive screening methods. We performed Spearman rank-order correlation to examine associations between primary care and gastroenterology physician supply and CRC screening prevalence, in terms of both overall CRC screening prevalence and prevalence of endoscopy, stool-based, and CT colonography, respectively. Analyses were conducted in SAS version 9.4 (SAS Institute Inc., Cary, NC). All P values were 2-sided, with *P*L<L.05 considered statistically significant.

## Results

The overall national prevalence of CRC screening for individuals aged 45-49 was 34.5% (95% CI: 33.4%–35.8%) in 2022. The prevalence of CRC screening was significantly higher among females than males (36.5%, 95% CI: 34.7%-38.4% vs. 32.7%, 95% CI: 31.1%-34.2%, *P* <.001). Other characteristics that were significantly associated with higher prevalence of CRC screening included non-Hispanic black race/ethnicity, being married, having a higher level of education, having an income of ≥$50,000, urban location, having insurance coverage, having a usual source of care, being a non-smoker or former smoker, being a non-drinker or casual drinker, being obese or overweight, having fair/poor self-rated general health, having two or more chronic conditions, and having two or more disabling limitations (**Table 1**). Prevalence of CRC screening varied significantly by region, with the highest prevalence occurring in the Northeast (35.9%, 95% CI: 33.3%-38.6%) and the lowest occurring in the West (31.9%, 95% CI: 29.3%-34.5%), as shown in **Table 2**. **Figure** 1 displays the prevalence of CRC screening across US states.

**Figure 1.**
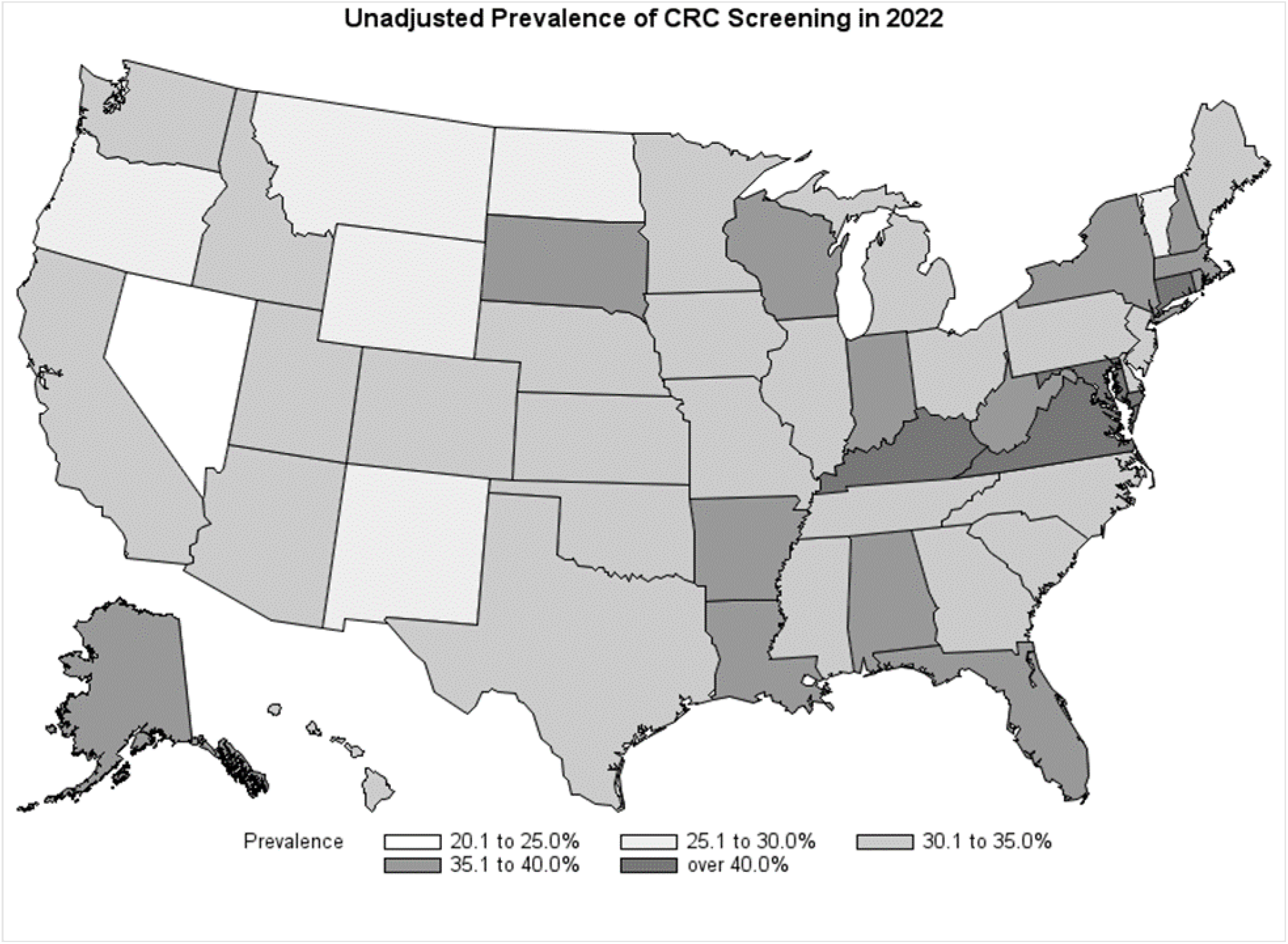
Map of CRC screening among Individuals Aged 45-49 by States in 2022.

**Table 1.** Prevalence of CRC Screening Uptake among Individuals Aged 45-49 in 2022, 2022 Adult Behavioral Risk Factor Surveillance System.

|  | Prevalence of CRC Screening % (95% CI) |  |  |  |  |  |  |  |  |  |  |  |  |  |  |  |  |  |  |  |
| --- | --- | --- | --- | --- | --- | --- | --- | --- | --- | --- | --- | --- | --- | --- | --- | --- | --- | --- | --- | --- |
|  | Overall |  |  | P-value | Northeast |  |  | P-value | Midwest |  |  | P-value | South |  |  | P-value | West |  |  | P-value |
|  | % | 95% CI |  |  | % | 95% CI |  |  | % | 95% CI |  |  | % | 95% CI |  |  | % | 95% CI |  |  |
| Sex |  |  |  | <.001 |  |  |  | 0.383 |  |  |  | 0.035 |  |  |  | 0.136 |  |  |  | 0.022 |
| Male | 32.7 | 31.1 | 34.2 |  | 34.8 | 31.4 | 38.2 |  | 32.2 | 29.4 | 34.9 |  | 34.2 | 31.4 | 37.0 |  | 28.9 | 25.7 | 32.0 |  |
| Female | 36.5 | 34.7 | 38.4 |  | 37.1 | 33.2 | 41.0 |  | 36.4 | 33.7 | 39.1 |  | 37.5 | 34.2 | 40.8 |  | 34.8 | 30.8 | 38.8 |  |
| Race/Ethnicity |  |  |  | <.001 |  |  |  | 0.292 |  |  |  | 0.980 |  |  |  | 0.002 |  |  |  | 0.048 |
| NH White | 35.1 | 33.9 | 36.3 |  | 36.3 | 33.2 | 39.4 |  | 34.6 | 32.5 | 36.7 |  | 36.1 | 33.9 | 38.2 | 0.000 | 32.9 | 30.1 | 35.6 |  |
| NH Black | 40.5 | 36.8 | 44.1 |  | 38.4 | 31.5 | 45.4 |  | 33.5 | 26.8 | 40.2 |  | 42.1 | 36.9 | 47.3 |  | 44.7 | 33.9 | 55.6 |  |
| NH Other | 29.6 | 25.6 | 33.7 |  | 28.7 | 19.5 | 38.0 |  | 33.4 | 24.6 | 42.1 |  | 23.4 | 16.7 | 30.2 |  | 32.6 | 25.4 | 39.8 |  |
| Hispanic | 32.1 | 28.4 | 35.7 |  | 38.1 | 31.0 | 45.1 |  | 33.7 | 26.5 | 40.9 |  | 33.6 | 27.1 | 40.2 |  | 27.2 | 21.5 | 33.0 |  |
| Marital Status |  |  |  | 0.013 |  |  |  | 0.022 |  |  |  | 0.134 |  |  |  | 0.602 |  |  |  | 0.017 |
| Married | 35.6 | 34.1 | 37.1 |  | 37.8 | 34.4 | 41.1 |  | 35.2 | 32.8 | 37.6 |  | 36.2 | 33.6 | 38.9 |  | 33.7 | 30.4 | 36.9 |  |
| Not Married | 32.4 | 30.3 | 34.4 |  | 32.0 | 28.3 | 35.7 |  | 32.2 | 29.1 | 35.3 |  | 35.0 | 31.2 | 38.8 |  | 27.5 | 23.7 | 31.3 |  |
| Education |  |  |  | <.001 |  |  |  | 0.019 |  |  |  | 0.002 |  |  |  | <.001 |  |  |  | 0.013 |
| Some or less than high school | 24.1 | 20.0 | 28.1 |  | 26.2 | 15.6 | 36.9 |  | 29.6 | 22.1 | 37.2 |  | 23.0 | 16.6 | 29.4 |  | 21.6 | 13.4 | 29.8 |  |
| High school graduate | 31.2 | 28.6 | 33.9 |  | 32.0 | 26.5 | 37.5 |  | 31.1 | 26.7 | 35.4 |  | 31.6 | 27.1 | 36.1 |  | 30.2 | 23.8 | 36.6 |  |
| Some college or technical school | 35.1 | 32.8 | 37.3 |  | 36.0 | 30.9 | 41.1 |  | 32.0 | 28.6 | 35.4 |  | 38.2 | 33.9 | 42.5 |  | 32.2 | 28.1 | 36.4 |  |
| College or technical school graduate | 39.7 | 38.0 | 41.5 |  | 40.4 | 36.9 | 44.0 |  | 39.7 | 36.9 | 42.5 |  | 41.7 | 38.6 | 44.8 |  | 36.3 | 32.3 | 40.3 |  |
| Income (\$) | | | | <.001 | | | | 0.067 | | | | 0.493 | | | | <.001 | | | | 0.220 |
| <15000 | 30.4 | 26.1 | 34.7 |  | 28.9 | 19.9 | 37.8 |  | 33.3 | 25.5 | 41.1 |  | 30.9 | 23.5 | 38.2 |  | 27.4 | 17.1 | 37.8 |  |
| 15000-<25000 | 29.2 | 25.2 | 33.3 |  | 32.1 | 23.0 | 41.2 |  | 34.1 | 26.7 | 41.6 |  | 29.6 | 23.0 | 36.1 |  | 23.5 | 15.0 | 32.0 |  |
| 25000-<35000 | 26.5 | 22.8 | 30.3 |  | 29.4 | 21.6 | 37.1 |  | 29.6 | 23.0 | 36.1 |  | 22.7 | 16.7 | 28.7 |  | 29.9 | 21.6 | 38.1 |  |
| 35000-<50000 | 32.4 | 28.2 | 36.5 |  | 34.5 | 21.5 | 47.5 |  | 31.8 | 25.2 | 38.5 |  | 34.7 | 28.6 | 40.7 |  | 27.2 | 18.0 | 36.5 |  |
| >=50000 | 37.7 | 36.1 | 39.3 |  | 39.4 | 36.0 | 42.8 |  | 35.5 | 33.0 | 38.0 |  | 40.3 | 37.3 | 43.3 |  | 34.5 | 31.3 | 37.7 |  |
| Unknown | 30.5 | 27.0 | 34.0 |  | 30.6 | 24.9 | 36.4 |  | 31.9 | 26.7 | 37.2 |  | 30.8 | 25.0 | 36.5 |  | 29.0 | 19.7 | 38.3 |  |
| Employment |  |  |  | 0.138 |  |  |  | 0.397 |  |  |  | 0.086 |  |  |  | 0.383 |  |  |  | 0.029 |
| Employed | 34.1 | 32.7 | 35.5 |  | 35.3 | 32.5 | 38.1 |  | 33.5 | 31.4 | 35.6 |  | 36.3 | 33.7 | 38.9 |  | 30.2 | 27.5 | 33.0 |  |
| Unemployed | 36.3 | 33.7 | 39.0 |  | 38.4 | 31.7 | 45.2 |  | 37.8 | 33.3 | 42.3 |  | 34.2 | 30.4 | 38.1 |  | 37.8 | 31.3 | 44.2 |  |
| **Urban/rural** |  |  |  | 0.029 |  |  |  | <.001 |  |  |  | 0.245 |  |  |  | 0.232 |  |  |  | 0.655 |
| Urban | 34.9 | 33.6 | 36.1 |  | 36.5 | 33.8 | 39.2 |  | 34.6 | 32.5 | 36.6 |  | 36.1 | 33.8 | 38.4 |  | 32.0 | 29.3 | 34.6 |  |
| Rural | 30.3 | 26.5 | 34.1 |  | 18.8 | 10.7 | 26.9 |  | 31.2 | 25.9 | 36.4 |  | 32.1 | 26.0 | 38.1 |  | 29.2 | 17.5 | 40.9 |  |
| **Health Insurance Coverage** |  |  |  | <.001 |  |  |  | 0.063 |  |  |  | <.001 |  |  |  | <.001 |  |  |  | <.001 |
| Private | 36.1 | 34.7 | 37.5 |  | 37.1 | 34.1 | 40.1 |  | 34.7 | 32.4 | 37.0 |  | 37.1 | 34.5 | 39.7 |  | 35.0 | 31.8 | 38.2 |  |
| Public | 38.6 | 36.0 | 41.1 |  | 38.2 | 32.8 | 43.5 |  | 39.1 | 35.0 | 43.3 |  | 44.0 | 39.6 | 48.4 |  | 30.4 | 25.0 | 35.7 |  |
| Uninsured | 19.8 | 15.8 | 23.8 |  | 24.7 | 13.7 | 35.8 |  | 20.4 | 13.9 | 26.8 |  | 19.7 | 13.2 | 26.2 |  | 15.9 | 9.8 | 22.0 |  |
| **Usual source of care** |  |  |  | <.001 |  |  |  | <.001 |  |  |  | <.001 |  |  |  | <.001 |  |  |  | <.001 |
| Yes | 37.6 | 36.2 | 38.9 |  | 38.2 | 35.4 | 41.0 |  | 36.9 | 34.7 | 39.0 |  | 39.2 | 36.8 | 41.7 |  | 35.0 | 32.1 | 38.0 |  |
| No | 18.5 | 16.0 | 21.0 |  | 19.2 | 13.6 | 24.7 |  | 18.5 | 15.2 | 21.8 |  | 19.5 | 15.0 | 24.1 |  | 16.5 | 12.2 | 20.7 |  |
| **Smoking Status** |  |  |  | 0.005 |  |  |  | 0.154 |  |  |  | 0.154 |  |  |  | 0.073 |  |  |  | 0.450 |
| Every day smoker | 29.9 | 26.8 | 33.0 |  | 30.2 | 22.4 | 37.9 |  | 32.0 | 27.3 | 36.7 |  | 30.5 | 25.3 | 35.7 |  | 25.6 | 18.2 | 32.9 |  |
| Someday smoker | 31.7 | 26.9 | 36.4 |  | 31.9 | 23.0 | 40.8 |  | 31.6 | 22.8 | 40.4 |  | 29.5 | 22.4 | 36.6 |  | 35.9 | 23.0 | 48.7 |  |
| Former smoker | 36.3 | 33.6 | 38.9 |  | 34.4 | 29.7 | 39.1 |  | 34.7 | 31.2 | 38.2 |  | 39.3 | 34.0 | 44.5 |  | 33.9 | 28.9 | 38.9 |  |
| Non-smoker | 34.9 | 33.3 | 36.5 |  | 37.7 | 34.1 | 41.3 |  | 34.3 | 31.6 | 37.1 |  | 35.9 | 33.2 | 38.6 |  | 31.8 | 28.4 | 35.3 |  |
| Unknown | 42.2 | 33.9 | 50.4 |  | 46.0 | 31.5 | 60.5 |  | 51.4 | 35.2 | 67.7 |  | 40.7 | 25.6 | 55.8 |  | 32.5 | 16.9 | 48.2 |  |
| **Drinking Status** |  |  |  | 0.018 |  |  |  | 0.162 |  |  |  | 0.323 |  |  |  | 0.323 |  |  |  | 0.212 |
| non-drinker | 33.5 | 31.7 | 35.2 |  | 35.0 | 30.7 | 39.3 |  | 33.9 | 30.9 | 36.9 |  | 34.1 | 31.2 | 37.0 |  | 31.1 | 27.0 | 35.2 |  |
| Casual drinker | 36.9 | 34.9 | 39.0 |  | 38.8 | 34.7 | 42.9 |  | 35.9 | 32.8 | 39.1 |  | 37.8 | 33.9 | 41.7 |  | 34.8 | 30.7 | 38.9 |  |
| Binge or heavy drinker | 33.0 | 30.3 | 35.7 |  | 32.5 | 27.5 | 37.6 |  | 32.0 | 28.0 | 36.1 |  | 36.4 | 31.0 | 41.8 |  | 28.7 | 23.4 | 34.0 |  |
| **BMI** |  |  |  | 0.003 |  |  |  | 0.316 |  |  |  | 0.007 |  |  |  | 0.028 |  |  |  | 0.368 |
| Lower-Normal | 33.6 | 31.0 | 36.2 |  | 36.3 | 31.4 | 41.2 |  | 33.8 | 29.6 | 38.0 |  | 34.0 | 29.3 | 38.7 |  | 31.1 | 25.4 | 36.8 |  |
| Overweight | 34.2 | 32.0 | 36.4 |  | 36.4 | 31.5 | 41.4 |  | 30.5 | 27.4 | 33.7 |  | 38.0 | 33.8 | 42.1 |  | 29.6 | 25.6 | 33.6 |  |
| Obesity | 36.8 | 34.9 | 38.6 |  | 37.4 | 33.1 | 41.6 |  | 37.8 | 34.7 | 41.0 |  | 36.8 | 33.6 | 39.9 |  | 35.2 | 31.0 | 39.4 |  |
| Unknown | 27.9 | 23.7 | 32.2 |  | 27.7 | 18.4 | 36.9 |  | 32.1 | 25.8 | 38.4 |  | 25.6 | 19.4 | 31.7 |  | 29.3 | 18.6 | 40.0 |  |
| **General Health** |  |  |  | 0.033 |  |  |  | 0.259 |  |  |  | 0.025 |  |  |  | 0.104 |  |  |  | 0.423 |
| Excellent | 31.8 | 29.0 | 34.5 |  | 32.4 | 27.4 | 37.3 |  | 27.9 | 23.2 | 32.6 |  | 32.8 | 27.8 | 37.7 |  | 32.7 | 26.5 | 38.8 |  |
| Very good | 34.2 | 32.2 | 36.2 |  | 36.9 | 32.5 | 41.4 |  | 35.1 | 31.7 | 38.4 |  | 35.3 | 31.7 | 39.0 |  | 29.6 | 25.4 | 33.8 |  |
| Good | 34.7 | 32.5 | 37.0 |  | 34.6 | 30.2 | 38.9 |  | 35.2 | 31.7 | 38.7 |  | 34.6 | 30.3 | 39.0 |  | 34.7 | 30.0 | 39.3 |  |
| Fair/poor | 37.8 | 34.8 | 40.7 |  | 40.6 | 32.7 | 48.5 |  | 37.5 | 33.1 | 41.8 |  | 41.0 | 36.3 | 45.7 |  | 30.0 | 23.5 | 36.5 |  |
| **# Chronic conditions** |  |  |  | <.001 |  |  |  | <.001 |  |  |  | <.001 |  |  |  | <.001 |  |  |  | <.001 |
| 0 | 30.2 | 28.6 | 31.7 |  | 32.3 | 29.2 | 35.3 |  | 29.4 | 27.0 | 31.8 |  | 31.9 | 29.1 | 34.7 |  | 26.5 | 23.5 | 29.6 |  |
| 1 | 38.2 | 35.8 | 40.7 |  | 39.3 | 33.7 | 45.0 |  | 38.8 | 34.9 | 42.7 |  | 38.3 | 33.7 | 42.9 |  | 36.9 | 32.1 | 41.7 |  |
| >=2 | 46.8 | 43.7 | 49.9 |  | 47.9 | 40.5 | 55.2 |  | 45.1 | 40.2 | 50.1 |  | 46.7 | 42.0 | 51.5 |  | 47.8 | 39.7 | 56.0 |  |
| **# Disabling limitations** |  |  |  | <.001 |  |  |  | 0.794 |  |  |  | <.001 |  |  |  | 0.102 |  |  |  | 0.135 |
| 0 | 33.3 | 31.9 | 34.7 |  | 35.8 | 32.9 | 38.8 |  | 32.3 | 30.1 | 34.5 |  | 34.6 | 32.0 | 37.2 |  | 30.4 | 27.6 | 33.2 |  |
| 1 | 37.5 | 34.0 | 41.0 |  | 38.0 | 30.4 | 45.6 |  | 38.2 | 33.1 | 43.4 |  | 37.4 | 31.5 | 43.4 |  | 36.6 | 28.3 | 44.9 |  |
| >=2 | 40.3 | 36.6 | 44.1 |  | 34.3 | 26.3 | 42.3 |  | 45.2 | 38.9 | 51.6 |  | 41.3 | 35.6 | 47.1 |  | 38.0 | 28.5 | 47.6 |  |

**Table 2.** State-level CRC Screening Prevalence among Individuals Aged 45-49 in 2022, 2022 Adult Behavioral Risk Factor Surveillance System.

|  | Unadjusted %<br>(95% CI) | 95% CI |  | Adjusted<br>Prevalence<br>Ratio* | 95% CI |  |
| --- | --- | --- | --- | --- | --- | --- |
| **National** | 34.5 | 33.4 | 35.8 |  |  |  |
| **Northeast** |  |  |  |  |  |  |
| Overall | 35.9 | 33.3 | 38.6 |  |  |  |
| Connecticut | 44.9 | 39.2 | 50.6 | 1.48 | 1.18 | 1.84 |
| Maine | 33.4 | 28.7 | 38.1 | 1.25 | 0.98 | 1.60 |
| Massachusetts | 36.4 | 31.7 | 41.2 | 1.23 | 0.98 | 1.55 |
| New Hampshire | 35.4 | 28.7 | 42.1 | 1.18 | 0.91 | 1.54 |
| Rhode Island | 35.4 | 28.2 | 42.6 | 1.20 | 0.92 | 1.57 |
| Vermont | 26.9 | 22.5 | 31.3 | Reference |  |  |
| New Jersey | 32.8 | 27.9 | 37.7 | 1.14 | 0.89 | 1.45 |
| New York | 38.8 | 34.8 | 42.9 | 1.35 | 1.09 | 1.68 |
| Pennsylvania | 32.5 | 24.4 | 40.7 | 1.18 | 0.90 | 1.55 |
| **Midwest** |  |  |  |  |  |  |
| Overall | 34.2 | 32.4 | 36.2 |  |  |  |
| Indiana | 36.1 | 31.9 | 40.3 | 1.21 | 0.95 | 1.55 |
| Illinois | 34.4 | 27.5 | 41.2 | 1.26 | 0.95 | 1.67 |
| Michigan | 33.9 | 29.3 | 38.5 | 1.12 | 0.87 | 1.45 |
| Ohio | 34.8 | 30.9 | 38.7 | 1.15 | 0.90 | 1.47 |
| Wisconsin | 39.3 | 34.7 | 44.0 | 1.26 | 0.98 | 1.61 |
| Iowa | 30.7 | 26.0 | 35.4 | 1.09 | 0.84 | 1.41 |
| Kansas | 33.1 | 28.3 | 37.9 | 1.17 | 0.91 | 1.51 |
| Minnesota | 31.1 | 27.6 | 34.6 | 1.12 | 0.88 | 1.42 |
| Missouri | 34.6 | 28.7 | 40.5 | 1.19 | 0.91 | 1.56 |
| Nebraska | 31.5 | 25.8 | 37.3 | 1.12 | 0.85 | 1.48 |
| North Dakota | 28.0 | 21.8 | 34.2 | Reference |  |  |
| South Dakota | 35.4 | 17.2 | 53.6 | 1.28 | 0.80 | 2.03 |
| **South** |  |  |  |  |  |  |
| Overall | 35.6 | 33.6 | 38.0 |  |  |  |
| Delaware | 32.8 | 24.7 | 41.0 | 0.94 | 0.68 | 1.28 |
| District of Columbia | 38.7 | 30.8 | 46.5 | 1.02 | 0.77 | 1.35 |
| Florida | 39.1 | 30.9 | 47.2 | 1.19 | 0.93 | 1.52 |
| Georgia | 32.1 | 26.2 | 38.1 | 0.89 | 0.68 | 1.17 |
| Maryland | 41.8 | 37.3 | 46.2 | 1.13 | 0.90 | 1.41 |
| North Carolina | 32.3 | 26.1 | 38.5 | 0.97 | 0.74 | 1.28 |
| South Carolina | 32.4 | 27.7 | 37.2 | 0.99 | 0.78 | 1.27 |
| Virginia | 40.1 | 35.0 | 45.2 | 1.13 | 0.90 | 1.42 |
| West Virginia | 38.3 | 32.2 | 44.3 | 1.12 | 0.87 | 1.43 |
| Alabama | 38.8 | 31.4 | 46.2 | 1.12 | 0.86 | 1.46 |
| Kentucky | 42.8 | 34.6 | 51.1 | 1.28 | 0.98 | 1.66 |
| Mississippi | 30.9 | 25.1 | 36.8 | 0.91 | 0.69 | 1.19 |
| Tennessee | 30.4 | 24.5 | 36.4 | Reference |  |  |
| Arkansas | 37.6 | 31.2 | 43.9 | 1.07 | 0.82 | 1.40 |
| Louisiana | 36.1 | 29.9 | 42.4 | 1.05 | 0.81 | 1.36 |
| Oklahoma | 32.8 | 27.5 | 38.1 | 1.00 | 0.76 | 1.31 |
| Texas | 34.4 | 29.2 | 39.7 | 1.08 | 0.85 | 1.36 |
| **West** |  |  |  |  |  |  |
| Overall | 31.9 | 29.3 | 34.5 |  |  |  |
| Arizona | 31.3 | 25.6 | 37.0 | 1.29 | 0.90 | 1.87 |
| Colorado | 31.9 | 28.0 | 35.8 | 1.22 | 0.85 | 1.74 |
| Idaho | 33.5 | 28.5 | 38.4 | 1.28 | 0.89 | 1.84 |
| New Mexico | 29.4 | 21.5 | 37.3 | 1.26 | 0.82 | 1.94 |
| Montana | 26.3 | 21.2 | 31.4 | 1.01 | 0.68 | 1.51 |
| Utah | 31.1 | 26.9 | 35.3 | 1.18 | 0.83 | 1.69 |
| Nevada | 24.6 | 16.1 | 33.2 | Reference | 0.66 | 1.47 |
| Wyoming | 29.2 | 22.6 | 35.8 | 1.11 | 0.72 | 1.69 |
| Alaska | 36.0 | 29.3 | 42.7 | 1.31 | 0.87 | 1.99 |
| California | 33.6 | 28.4 | 38.8 | 1.39 | 0.98 | 1.97 |
| Hawaii | 32.0 | 26.2 | 37.9 | 1.22 | 0.83 | 1.81 |
| Oregon | 27.5 | 22.5 | 32.6 | 1.05 | 0.72 | 1.53 |
| Washington | 31.4 | 28.8 | 34.0 | 1.17 | 0.83 | 1.64 |

\* Adjusted for sex, race/ethnicity, marital status, education, income, employment, urban/rural, health insurance coverage, usual source of care, smoking, drinking, BMI, general health, comorbidity, and number of disabling limitations.

As shown in **Table 3**, among those who underwent a single type of screening, Endoscopic tests were the most common screening method (74.9%), followed by stool-based tests (9.3%) and CT colonography (0.5%). Combined methods including endoscopy were more prevalent (9.6%) than those without (5.6%). Overall, 15.4% chose non-invasive screening. Significant differences in screening preferences were observed based on region, gender, race/ethnicity, insurance status, education, and income. The West showed the highest preference for non-invasive methods (23.8%, P<.001). Males, non-Hispanic other and Hispanic individuals, uninsured people, and those with lower education and income levels also preferred non-invasive methods more than their counterparts. **Figure 2** depicts bar charts of the patterns we observed that were statistically significant, while the bar chart for the non-significant association (urban/rural location) is included in the Supplemental Materials.

**Figure 2.**
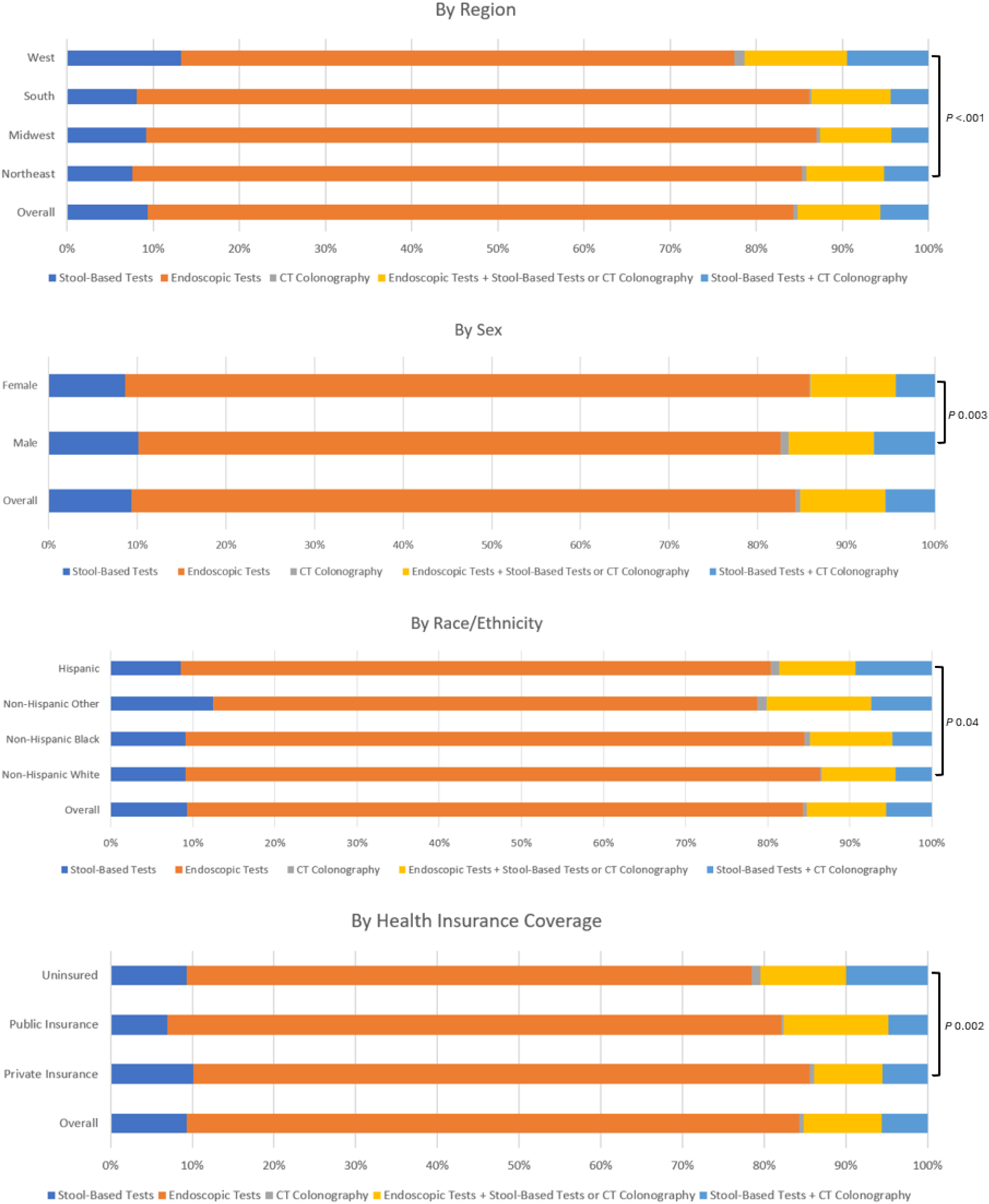
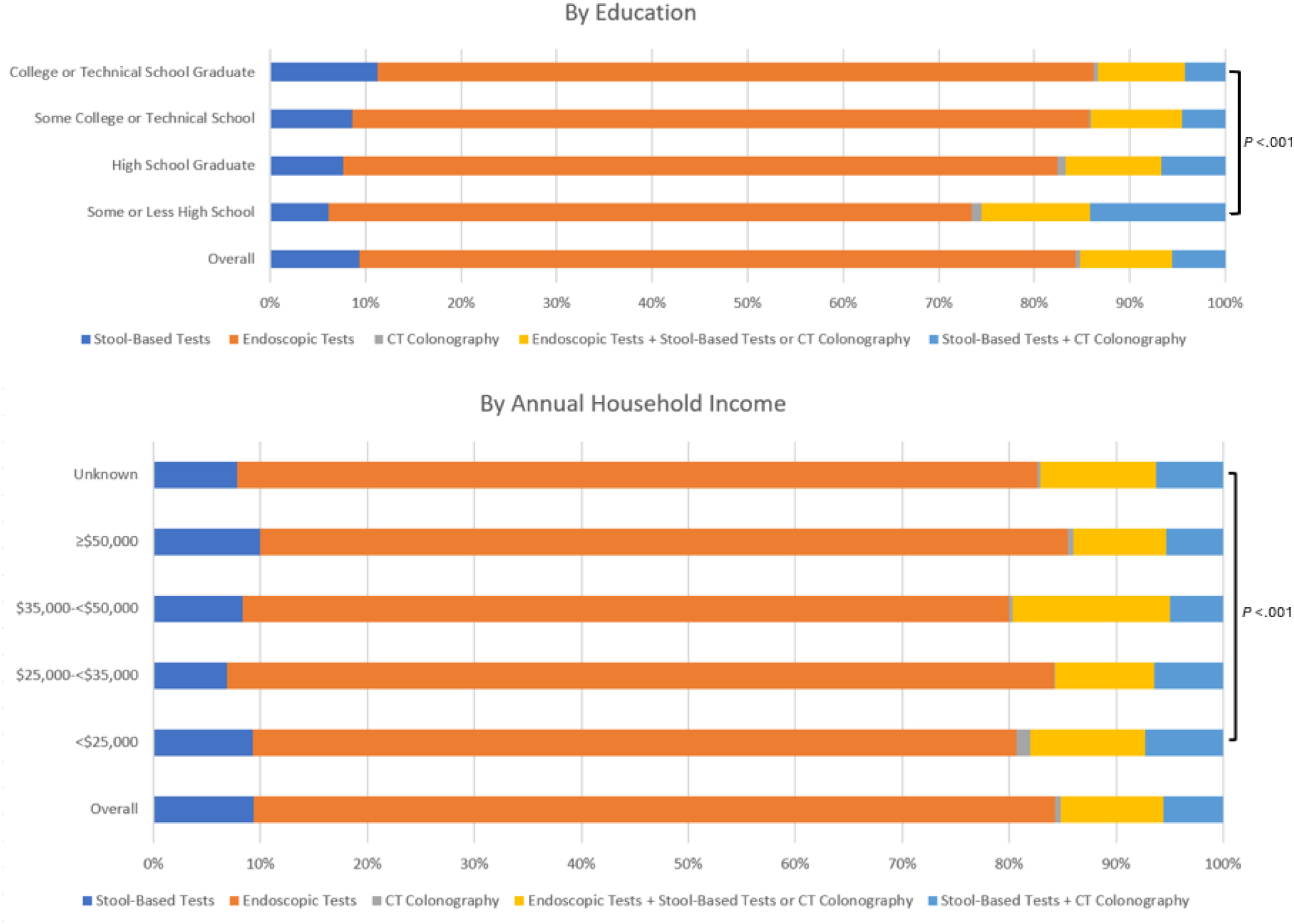
Bar Charts of Types of Screening Modality Undergone, Overall and By Region, Sex, Race/Ethnicity, Health Insurance Coverage, Education, and Annual Household Income, 2022 Adult Behavioral Risk Factor Surveillance System.

**Table 3.** Prevalence of Screening by Type of Screening Modality According to Sociodemographic Characteristics.

|  | Type of Screening Modality |  |  |  |  | P-Value |
| --- | --- | --- | --- | --- | --- | --- |
|  | Stool-Based Tests (%) | Endoscopic Tests (%) | CT colonography (%) | Combination of Endoscopic Tests and Stool-Based Tests or CT Colonography (%) | Combination of Stool-Based Tests and CT Colonography (%) |  |
| **Overall** | 9.3 | 74.9 | 0.5 | 9.6 | 5.6 |  |
| **Region** |  |  |  |  |  | <.001 |
| Northeast | 7.6 | 77.7 | 0.5 | 9.0 | 5.2 |  |
| Midwest | 9.2 | 77.8 | 0.4 | 8.3 | 4.3 |  |
| South | 8.1 | 78.1 | 0.3 | 9.2 | 4.4 |  |
| West | 13.2 | 64.2 | 1.1 | 11.9 | 9.5 |  |
| **Sex** |  |  |  |  |  | 0.003 |
| Male | 10.1 | 72.4 | 0.9 | 9.6 | 6.9 |  |
| Female | 8.6 | 77.2 | 0.2 | 9.6 | 4.4 |  |
| **Race/Ethnicity** |  |  |  |  |  | 0.04 |
| Non-Hispanic White | 9.1 | 77.3 | 0.2 | 9.0 | 4.4 |  |
| Non-Hispanic Black | 9.1 | 75.4 | 0.6 | 10.1 | 4.8 |  |
| Non-Hispanic Other | 12.5 | 66.3 | 1.2 | 12.7 | 7.4 |  |
| Hispanic | 8.6 | 71.8 | 1.0 | 9.3 | 9.3 |  |
| **Health Insurance Coverage** |  |  |  |  |  | 0.002 |
| Private Insurance | 10.1 | 75.5 | 0.5 | 8.4 | 5.5 |  |
| Public Insurance | 7.0 | 75.1 | 0.3 | 12.8 | 4.8 |  |
| Uninsured | 9.3 | 69.2 | 1.0 | 10.5 | 10.0 |  |
| **Urban/Rural** |  |  |  |  |  | 0.25 |
| Urban | 9.4 | 75.1 | 0.5 | 9.5 | 5.4 |  |
| Rural | 7.3 | 71.1 | 0.4 | 11.2 | 10.1 |  |
| **Education** |  |  |  |  |  | <.001 |
| Some or less than high school | 6.1 | 67.3 | 1.1 | 11.3 | 14.2 |  |
| High school graduate | 7.6 | 74.8 | 0.9 | 10.0 | 6.7 |  |
| Some College or Technical School | 8.6 | 77.2 | 0.1 | 9.6 | 4.5 |  |
| College or Technical School Graduate | 11.3 | 75.0 | 0.5 | 9.1 | 4.2 |  |
| **Income** |  |  |  |  |  | <.001 |
| <\$25,000 | 9.3 | 71.4 | 1.3 | 10.7 | 7.3 | |
| \$25,000-<\$35,000 | 6.9 | 77.3 | 0. | 9.3 | 6.4 | |
| \$35,000-<\$50,000 | 8.3 | 71.7 | 0.3 | 14.7 | 5.0 | |
| ≥\$50,000 | 9.9 | 75.6 | 0.5 | 8.7 | 5.3 | |
| Unknown | 7.8 | 74.9 | 0.2 | 10.8 | 6.3 |  |

**Figure 3** shows significant correlation scatterplots between gastroenterology physician supply and both overall CRC screening prevalence and endoscopy screening prevalence. Non-significant relationships are in the **Supplemental Materials**. Physician supply varied widely across the US, with the District of Columbia having the highest supply of both primary care (504.9) and gastroenterology physicians (28.7) per 10,000 population aged 45-49 years. Mississippi and Alaska had the lowest supply of primary care (104.7) and gastroenterology physicians (3.1), respectively. Gastroenterology physician supply positively correlated with overall CRC screening prevalence (Spearman’s ρ=0.42, P=.002) and endoscopy screening prevalence (ρ=0.39, P=.005). Other physician supply and screening type combinations showed positive but non-significant associations. CT colonography screening facilities ranged from 0 in five states to 27 in California, New York, and Florida. The number of facilities weakly correlated with CT colonography screening prevalence (ρ=0.18, P=.22) (**Supplemental Materials**).

**Figure 3.**
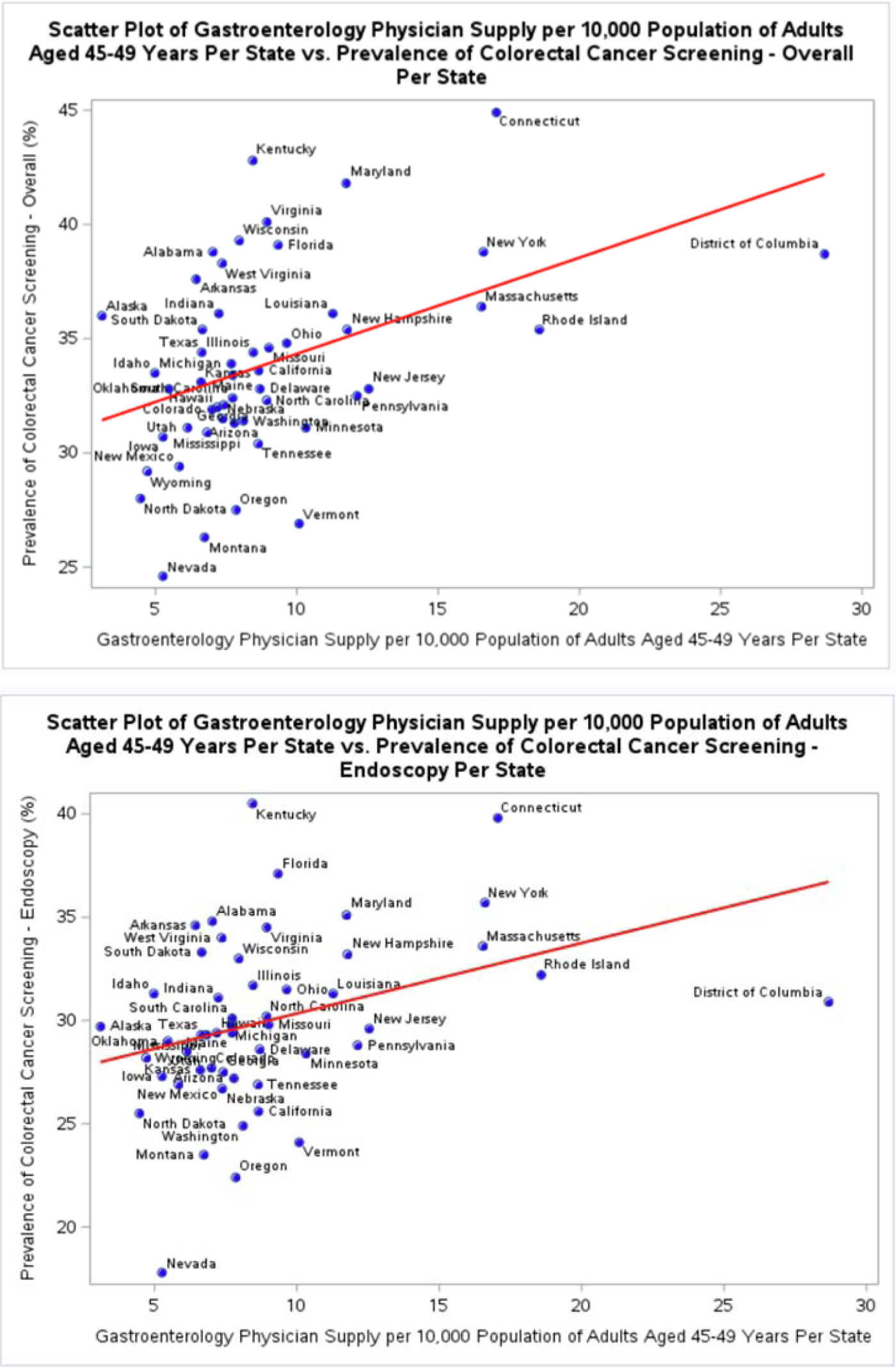
Correlation Scatter Plots of Overall CRC Screening Prevalence and Endoscopy Screening Prevalence According to Supply of Gastroenterology Physicians by State, 2022 Adult Behavioral Risk Factor Surveillance System. Moderate significant positive associations were detected between supply of gastroenterology physicians per state and overall prevalence of CRC screening (Spearman’s ρ 0.42, *P* 0.002) and prevalence of endoscopy (Spearman’s ρ 0.39, *P* 0.005).

## Discussion

In this analysis of nationally representative data from the 2022 BRFSS, we observed low CRC screening rates among adults aged 45-49 years. Despite the USPSTF’s recommendation in 2021 that average-risk adults in this age group undergo screening,^1^ only just over one-third of them in 2022 reported having undergone any modality of CRC screening. This figure is lower than expected; by comparison, a prior study that examined data from the National Health Interview Survey (NHIS) reported that, in 2018, overall CRC screening prevalence was 66.1% among all age groups, and 47.6% for 50–54-year-olds – the youngest age group at that time for which screening was recommended.^6^ These figures are higher than the 34.5% prevalence rate of CRC screening that we observed among 45–49-year-olds in our study. However, it is important to note that the guideline change is relatively recent, and it may take time for screening rates to increase in this newly eligible age group.

We also examined patterns in CRC screening modalities among this newly eligible 45-49 age group. Overall, endoscopic tests were the most predominant screening method. The high prevalence of endoscopic screening among this young adult population, in comparison to other screening modalities, may be partly due to the referral of individuals with positive results from non-invasive tests to diagnostic colonoscopies.^49–52^ Furthermore, a lack of widespread awareness about CRC screening guidelines in this younger cohort may result in symptomatic individuals being directly referred for colonoscopy during routine check-ups or emergency department visits, potentially inflating the prevalence of invasive screening methods in comparison to non-invasive methods.^53–55^

Of note, we observed significant variations in the types of screening modality undergone according to sociodemographic factors. Lower socioeconomic status and lack of health insurance were both associated with a greater preference for non-invasive screening methods. These findings are consistent with those noted in previous studies.^15,56,57^ The lower uptake of invasive screening methods observed among those with lower levels of both income and education, among those without health insurance coverage, and among non-Hispanic other and Hispanic individuals, suggest that persistent disparities in healthcare access may affect individuals’ choice of screening modality. Furthermore, cost may play a significant role; one study estimated that the average cost of a colonoscopy was $1,425 compared to $1,018 for CT colonography among a commercially insured population,^58^ while another study found, for Medicare, the average cost of a colonoscopy procedure was estimated to be $1,035 compared to $439 for a CT colonography.^59^ This suggests that individuals of disadvantaged socioeconomic status might face cost barriers to endoscopic tests resulting in greater preference for non-invasive methods among these groups. Taken together, our findings further support the need for a multifaceted approach to improve CRC screening rates and reduce disparities in this newly eligible population.^60–62^ Future interventions should focus on developing targeted, culturally sensitive educational campaigns to increase awareness of all CRC screening options and investigate the decision-making processes of both patients and healthcare providers when selecting screening modalities. Additionally, further research with a qualitative approach is warranted to elucidate the interplay between this population’s socioeconomic factors, healthcare access, and screening preferences.

The low overall CRC screening rate we observed in this study may be partly due to a time lag; given that the new USPTF screening recommendation went into effect in May 2021, it is possible that not enough time had passed for a strong effect to be seen in 2022 data. Furthermore, the impact of the COVID-19 pandemic may have resulted in delayed or cancelled screenings.^39^ Thus, future research should continue to monitor trends in CRC screening rates in this age group to assess whether screening rates are increasing over time. However, it is also worth noting that the American Cancer Society made a similar recommendation in 2018 that average-risk adults should begin CRC screening starting at age 45.^40^ Our findings, therefore, suggest there may be other underlying reasons for the continued low screening uptake among this age group. Another possible explanation for low CRC screening uptake among this age group is competing demands and priorities, such as work and family and caregiving responsibilities.^6,8,41,42^ Those in the 45-49 years age group, being ineligible for Medicare, may also be less likely to have adequate insurance coverage, which is another barrier to screening.^43^ Additional reasons for low CRC screening uptake in this age group include lower perceived risk with cancer screening not being prioritized,^7^ and reduced engagement with primary care due to lower prevalence of chronic conditions in this age group compared with older adults.^6,11,44,45^ However, colorectal cancer incidence is increasing among younger adults,^46,47^ highlighting the need for targeted efforts to increase CRC screening uptake among this group. We recommend that providers initiate early discussions about the importance of CRC screening with their patients before age 45, to ensure that screening begins at the recommended age.^30^ Furthermore, patients should be informed about the availability of multiple screening options including take-home options.^48^

We also observed a significant association between gastroenterology physician supply and both overall screening prevalence and prevalence of endoscopy screening. This association is supported by a 2017 Medicare claims data study, which showed that gastroenterologists performed the majority (57.2%) of colonoscopies.^35^ Our findings suggest that individuals who live in areas underserved by gastroenterology physicians may have reduced access to endoscopy screening procedures. This may partly explain why we observed that individuals in rural areas reported lower CRC screening rates than those in urban areas, since previous data has shown that the density of gastroenterologists is higher in urban areas.^36^ Furthermore, in “Looking at the Future of Gastroenterology”, the American Gastroenterology Association noted that the gastroenterologist workforce has not increased in line with the increasing population of the US and increasing gastroenterological disease burden.^37,38^ A follow-up modelling study predicted a future shortage of gastroenterologists and hepatologists.^37,38^ Our findings emphasize the importance of addressing these projected shortages in the gastroenterology physician workforce.

Our study has several strengths, including our use of nationally representative data. Our large sample size enabled us to estimate the prevalence of CRC screening overall, by test type, among geographic regions and within population subgroups. Furthermore, our inclusion of state-level data from the AHRF, and information on the locations of CT colonoscopy facilities from the ACR, allowed us to analyze associations between CRC screening and primary care physician supply, gastroenterology physician supply, and CT colonoscopy screening facility availability. Our study also has some limitations. The information in the BRFSS was self-reported and thus some respondents may have incorrectly reported their CRC screening history. Some studies have found that individuals tend to over-report their CRC screening history,^63,64^ while other studies have found relatively good corroboration between self-reported CRC screening use and that which is reported in medical records.^65–68^ Another limitation was that the ACR was only able to provide us with data on the locations of ACR accredited CT colonoscopy screening facilities, thus it is possible that other non-ACR accredited CT colonoscopy facilities were not accounted for in our analyses.

## Conclusion

Following the USPTF’s 2021 recommendation for average-risk adults aged 45-49 years to undergo CRC screening, screening rates are suboptimal, with just over one-third of adults in this age group reporting having undergone any modality of CRC screening. Disparities were observed according to geographic region and sociodemographic factors. Furthermore, individuals of lower socioeconomic status exhibited a greater preference for non-invasive screening modalities, which may be indicative of disparities in access to endoscopy. Significant associations were observed between gastroenterology physician supply and prevalence of both overall CRC screening and endoscopy screening, highlighting the importance of addressing projected shortages in the gastroenterology physician workforce. Our findings emphasize the need for targeted efforts to increase CRC screening uptake among adults aged 45-49 years, with a focus on ensuring equitable access to health-promoting information and screening services among underserved populations to prevent the widening of health disparities by SES and racial/ethnic groups.

## Data Availability

The data used in this study are publicly available from the Centers for Disease Control and Prevention (CDC). The Behavioral Risk Factor Surveillance System (BRFSS) 2022 dataset can be accessed through the CDC’s official BRFSS website: https://www.cdc.gov/brfss/annual_data/annual_2022.html. The Area Health Resources Files (AHRF) are available from the Health Resources and Services Administration (HRSA) Data Warehouse: https://data.hrsa.gov/topics/health-workforce/ahrf. The list of American College of Radiology (ACR)-accredited CT colonography screening facilities was obtained from the ACR and is publicly accessible through their website: https://www.acr.org/Clinical-Resources/Colon-Cancer-Screening-Resources/My-CT-Colonography.

## Author contributions

Rachel Liu-Galvin, MBChB (Data curation; Formal analysis; Investigation; Methodology; Visualization; Writing - original draft; Writing - review & editing), Zhigang Xie, PhD, B. Med, MPA (Conceptualization, Data curation; Software; Formal analysis; Investigation; Methodology; Visualization; Writing - original draft; Writing - review & editing), Young-Rock Hong, PhD, MPH (Conceptualization, Data curation; Investigation; Methodology; Writing - original draft; Writing - review & editing; Project administration; Supervision)

## Funding

None

## Conflicts of interest

The authors declare no conflict of interest.

## Acknowledgements

We are grateful to the American College of Radiology for providing us with a list of all ACR-accredited CT colonography screening facilities in the US as of July 22^nd^, 2024.

## Notes

### Competing Interest Statement

The authors have declared no competing interest.

### Funding Statement

This study did not receive any funding.

### Author Declarations

The study used ONLY openly available human data that were originally located at: the Behavioral Risk Factor Surveillance System (BRFSS) 2022 dataset: https://www.cdc.gov/brfss/annual_data/annual_2022.html

